# Catalysing Interprofessional Eye Health Education Using the Arclight diagnostic tool and simulation eyes in Rwanda: Outcomes from a Mixed Methods Randomised Trial

**DOI:** 10.64898/2026.02.12.26346142

**Authors:** Gatera Fiston Kitema, Veronica O’Carroll, Anita Laidlaw, Jean Baptiste Sagahutu, Andrew Blaikie

**Affiliations:** School of Health Sciences, College of Medicine and Health Sciences, University of Rwanda, Kigali, Rwanda; School of Medicine, University of St-Andrews, Scotland, United Kingdom; School of Medicine, Medical Sciences and Nutrition, University of Aberdeen, Scotland, United Kingdom

**Keywords:** Interprofessional Education, Visual Impairment, Arclight, Sub-Saharan Africa, Primary Eye care, Collaborative Eye Health

## Abstract

**Background:** Vision loss represents a significant public health concern according to the World Health Organisation, with increasing global age-standardised prevalence rates. Visual impairments are disproportionately distributed, occurring eight times more frequently in Sub-Saharan Africa and Southeast Asia compared to high-income regions. The Interprofessional Education (IPE) approach, utilizing the Arclight package as an implementation vehicle, offers promising potential for collaborative early detection and management of eye conditions in resource-constrained environments. This research aimed to implement validated Interprofessional Eye Health Education(IPEHE) in Rwanda, assess fundamental eye health knowledge and skills acquisition, evaluate medium to long-term learning retention, and explore IPE’s role in developing these competencies.

**Methods:** The study employed a mixed-methods approach combining a Randomised Controlled Trial (RCT) with qualitative assessment at the University of Rwanda. Researchers invited 443 final-year students from diverse healthcare programs including nursing, pharmacy, midwifery, medicine, and ophthalmic clinical officers. With statistical power set at 0.80 and alpha error probability at 0.05, the design aimed to detect pre-post training score differences of 10% or greater. The calculated sample size of 54 participants per group was expanded to 280 total participants (180 intervention, 100 control) to accommodate potential attrition.

**Results:** In the intervention group, 161 students (89.4%) attended the training, and 113 (70.2%) participated in the 10-month follow-up assessment (POST2). Of the control group, 90 participants (90%) attended assessments at 10 months post-intervention (POST2). Knowledge scores in the intervention group increased by 58.9% (SD=20.8, Z=10.82 p<0.001) immediately post-training, while skills improved by 49.7% (SD=14, Z=−8.382, P<0.001). At the 10-month follow-up, these gains remained significantly above baseline levels. Intervention participants significantly outperformed the control group at follow-up in both knowledge, with a 54.1% difference (SE= 2.0, df (201) = 27.3, P<0.001), and skills, with a 44% difference (SE=1.1, t(155)=38.7, p<0.001). Qualitative data from the intervention group indicated an appreciation for interprofessional collaboration, holistic patient care approaches, and the practical skills acquired.

**Conclusion:** The IPEHE intervention significantly enhanced collaborative eye health knowledge and skills among Rwandan healthcare students, with demonstrated retention up to 10 months post-intervention. These findings suggest that pre-qualification interprofessional education effectively produces collaborative practice-ready professionals capable of addressing eye health challenges in resource-limited settings.

## Introduction

Visual impairment is over eight times more prevalent in Sub-Saharan Africa (SSA) and South Asia compared to high-income regions.^(1)^ In SSA, 111 million people are affected by vision loss, with an age-standardised prevalence of over 18%^(2)^ and the burden continues to rise^(1)^.There is consequently an urgent call to action to tackle growing visual impairments in developing countries using the limited existing services in a cost-effective way.^(2)^ Rwanda like other low income countries experiences a significant and steady increase in demand for eye health services.^(3)^ It has been suggested that around one-third of this burden could be addressed through the delivery of more integrated primary eye care (PEC) services.^(4)^

Although various initiatives have made a positive impact on blindness reduction, the need is clearly still greatest where access to eye health specialists and diagnostic equipment is least^(5)^ with low and middle income countries (LMICs) disproportionately impacted. This is in large part due to a chronic shortage of equipment, training and trained eye care specialists in South Asia and sub-Saharan Africa (SSA).^(6)^

The Vision 2020 “The Right to Sight” initiative aimed to reduce avoidable blindness worldwide.^(6)^ A key strategy was the development of mid-level eye care professionals, such as ophthalmic clinical officers (OCOs) and ophthalmic nurses (OPNs), to support ophthalmologists in diagnosing and managing common eye conditions at the district hospital level. While this has helped address workforce shortages at secondary care level, there remains a pressing need to strengthen collaboration and integration of services at the community level.

The World Health Organization (WHO) is now increasingly advocating for collaborative approaches to tackle global health challenges sustainably.^(7)^ In eye health, this means integrating services within broader health systems and encouraging teamwork across professional boundaries.^(2)^

Interprofessional education (IPE), defined as “occasions when members or students of two or more professions learn with, from, and about each other to improve collaboration and the quality of care and services”^(8)^, offers a promising pathway to build this collaborative practice and enhance the effectiveness of eye care delivery. In the SSA context, evidence suggests that IPE can improve collaborative practice through the understanding of each profession’s role and the need for a holistic approach to health care.^(9)(10)(11)^ Several collaborative practice programs for eye health have been implemented in SSA to enhance the effectiveness of eye care delivery. For instance, Rwanda has attempted to integrate Primary Eye Care services into health centers close to the community, and nurses now assess and manage minor eye diseases and uncorrected refractive errors such as presbyopia.^(12)^ In this case of collaborative practice, more serious and complex cases are referred to secondary and tertiary-level national units where ophthalmic clinical officers have been trained to provide higher-level eye care services.^(13)^ However, this collaboration is more role and responsibility-oriented than it is an actual interprofessional collaboration. IPE has been identified as a way of improving interprofessional collaborative practice between professionals and at pre-qualification level, preparing students or trainees for future collaborative practice.^(14)^

A pilot study conducted at the University of Rwanda by O’Carroll et al^(6), (15)^ indicated that the successful use of IPE to deliver eye care knowledge and skills is feasible with the use of frugal diagnostic tools such as the Arclight. Whilst the findings from that pilot indicated some impact of IPE on students’ learning, such as students’ acquisition of fundamental eye health knowledge and skills, it did not follow up on the mid to long-term retention of this knowledge and these skills. This study sought to implement validated interprofessional eye health education (IPEHE) in Rwanda and explore the gap in evidence on interprofessional eye health knowledge and skills long term retention following such training. This would be with the long-term goal of establishing more effective community level eye care to reduce avoidable blindness consistent with current supranational recommendations.^(2)^

This study consequently aimed to answer the following questions:

1. Does the IPEHE intervention improve fundamental eye health knowledge in the short and longer term?
2. Does the IPEHE intervention improve fundamental eye health skills in the short and longer term?
3. Does the IPE approach contribute to collaborative eye health knowledge and skills acquisition through the IPEHE intervention?

## Methods

An explanatory sequential mixed-methods randomised study^(16)^ was conducted to answer the research questions. A randomised controlled trial (RCT) component explored “what” and “how much” participants were able to learn. An exploratory qualitative component indicated “How” participants acquired knowledge and skills from the training program and approach.

This study was conducted at the University of Rwanda with final-year students. An interprofessional sample of students grouped according to their professions was randomised to either attend the intervention (IPEHE) or control. Qualitative feedback related to the training was collected from IPEHE participants.

### Participants and recruitment

The study population was all 443 final-year undergraduate students from five disciplines: nursing, general medicine, midwifery, pharmacy, and ophthalmic clinical officers. These professions were selected as they represent the disciplines that deliver the majority of health care in Rwanda and are also involved in managing many eye disorders.^(12)^ The selected professions were also chosen based on being among frontline health workers who encounter patients, including those with eye problems.^(17)^

The sample size calculation approach was underpinned by CONSORT^(18),(19)^ guidelines whereby the power was set at 0.80 and the probability of Alpha error at 0.05. The study was designed to identify a difference in pre and post-training scores of 10% and above based on the previous study by O’Carroll and colleagues^(6),(15)^. A pilot study conducted to validate a questionnaire assessment tool identified a mean difference of almost five times the effect size with a standard deviation of around 10%. Using G-power software^(20)^, the calculated sample size was 54 participants in each group. The sample size was increased to 280 with 180 participants in the intervention group and 100 in the control group to mitigate effects from loss to follow-up and participant dropout.

Participants were invited through faculty and class representatives to participate in this study. Participants confirmed their willingness to participate by signing the consent form after reading the participant information sheet before randomisation. All non-ophthalmology students were asked whether they previously attended or participated in any form of ophthalmology training apart from what is usually covered in the teaching curriculum at the University of Rwanda, College of Medicine and Health Sciences(UR-CMHS). The recruitment of participants took place in the period between 21^st^ September 2021 to 30^th^ December 2021.

Block randomisation^(21)^ was applied to ensure there was no difference in distribution by gender and discipline between the intervention and control groups using an online tool.^(22)^ Randomisation was masked to participants prior to the intervention to avoid bias. All participants other than ophthalmic clinical officers reported no prior ophthalmology training or workshops (except what is covered in their respective curricula).^(22)^

### Intervention description

The IPEHE training package was designed and delivered with the intention that knowledge, skills, and behaviour could be transferred into practice according to the transfer of training theory adapted by Grossman and Salas.(^238)^ The intervention addressed both theoretical knowledge and practical skills that were acquired collaboratively following IPE principles of ensuring that learning was active and interactive amongst the students^(23)^ to enable learning with, from and about^(8)^. As part of theoretical knowledge, participants covered: interprofessional education and collaborative practice, biopsychosocial approach in health care delivery, the importance of primary eye care, normal eye anatomy and visual function, signs and symptoms of common eye diseases that cause red eye and loss of vision. The skills training included: holistic history taking, visual acuity testing for distance and near, dispensing near vision glasses, pupil examination, fundal reflex examination, essentials of anterior segment examination, fundamentals of eye care, counselling and appropriate referral and interprofessional practice.

### The Arclight package and simulation eyes

The Arclight package and simulation eyes were central to the delivery of the intervention. These low-cost diagnostic and training simulation tools were developed by the University of St Andrews in collaboration with Arclight Medical Ltd to improve training, equipping and access to eye care and has been shown to be an effective tool in LMICs as well as HICs.^(24)^ The package includes a cloth visual acuity chart, a reading chart, a lanyard, a mobile phone camera clip, a USB-C cable, a monster to help with fixation when examining babies and the Arclight device (anterior segment loupe, direct ophthalmoscope, and otoscope), all included in a small fabric case.(Figure 1) Simulation eyes for teaching to simulate different eye conditions ranging from leukocoria to different retinal conditions and have also been shown to be an effective training tool.^(25)^

**Figure 1:**
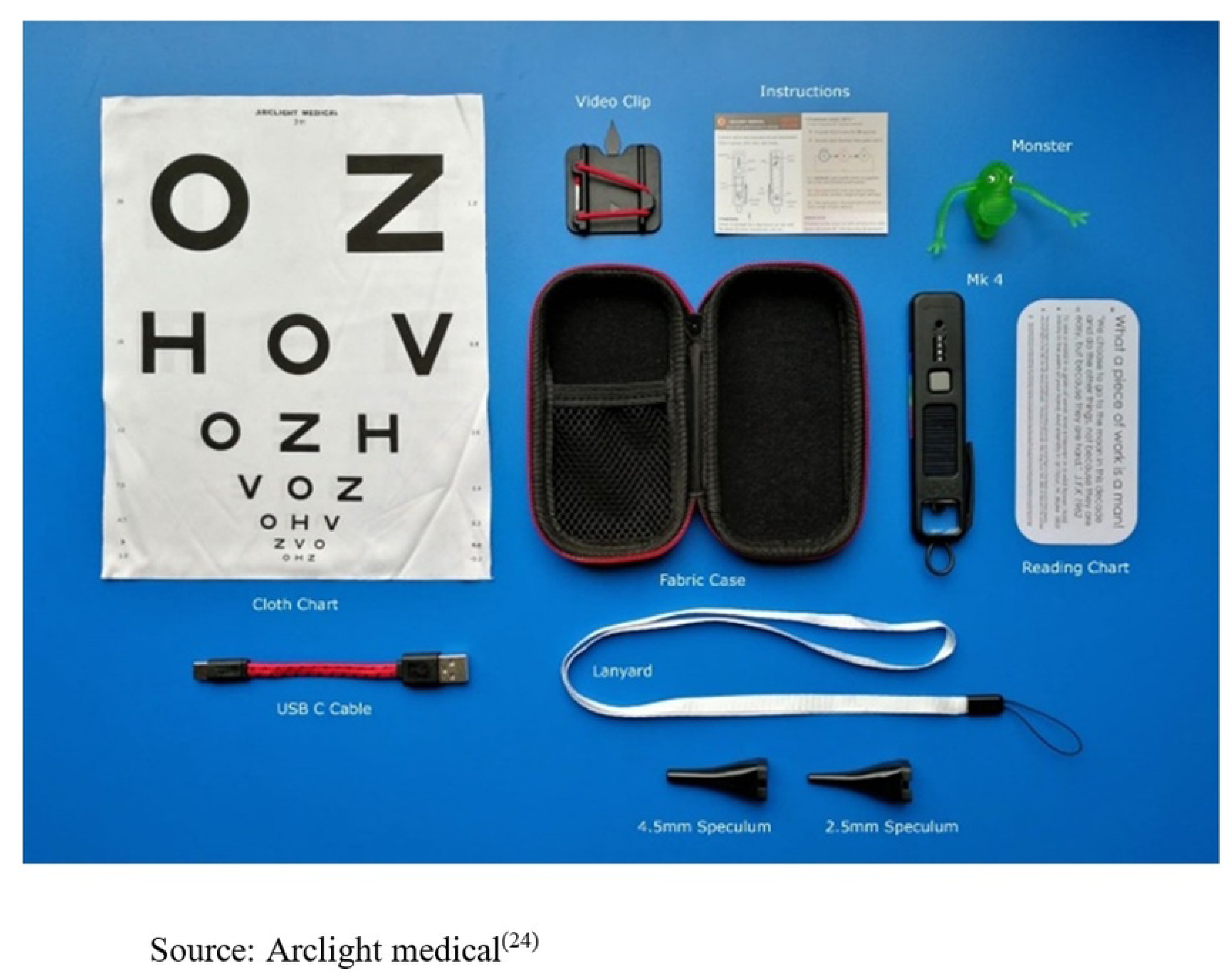
Photo of the Arclight ophthalmoscope with accessories.

Importantly, all these devices are low-cost, highly portable, and powered by a solar panel and rechargeable lithium battery combination that illuminates a Light-Emitting Diode (LED) light source.^(26),(27)^

Previous studies in Rwanda^(6),(15)^ suggested that using the Arclight and the IPE approach, different professions like nurses, clinical officers, medical doctors, and ophthalmic clinical officers can acquire immediate post training knowledge, skills, and confidence in identifying fundamental eye conditions in line with the WHO PEC manual^(28)^.

The validated intervention comprised two days of in-person training, whilst the control group were given similar course content to review individually.

### Outcome Assessment

The Kirkpatrick model adapted by Barr^(29)^ for training evaluation, a framework for outcome assessment was implemented. Knowledge and skills were assessed before (pre-training assessment – PRE) the training to determine the baseline, and a similar assessment was conducted immediately after the training (post-training assessment – POST1) and 10 months later to assess learning retention (POST2). At 10 months, collaborative eye health knowledge and skills were compared between the intervention and control groups. Due to resource constraints, only the intervention group attended pre- and immediate post-assessments, and the control group was assessed only 10 months after the intervention (see Figure 2).

**Figure 2:**
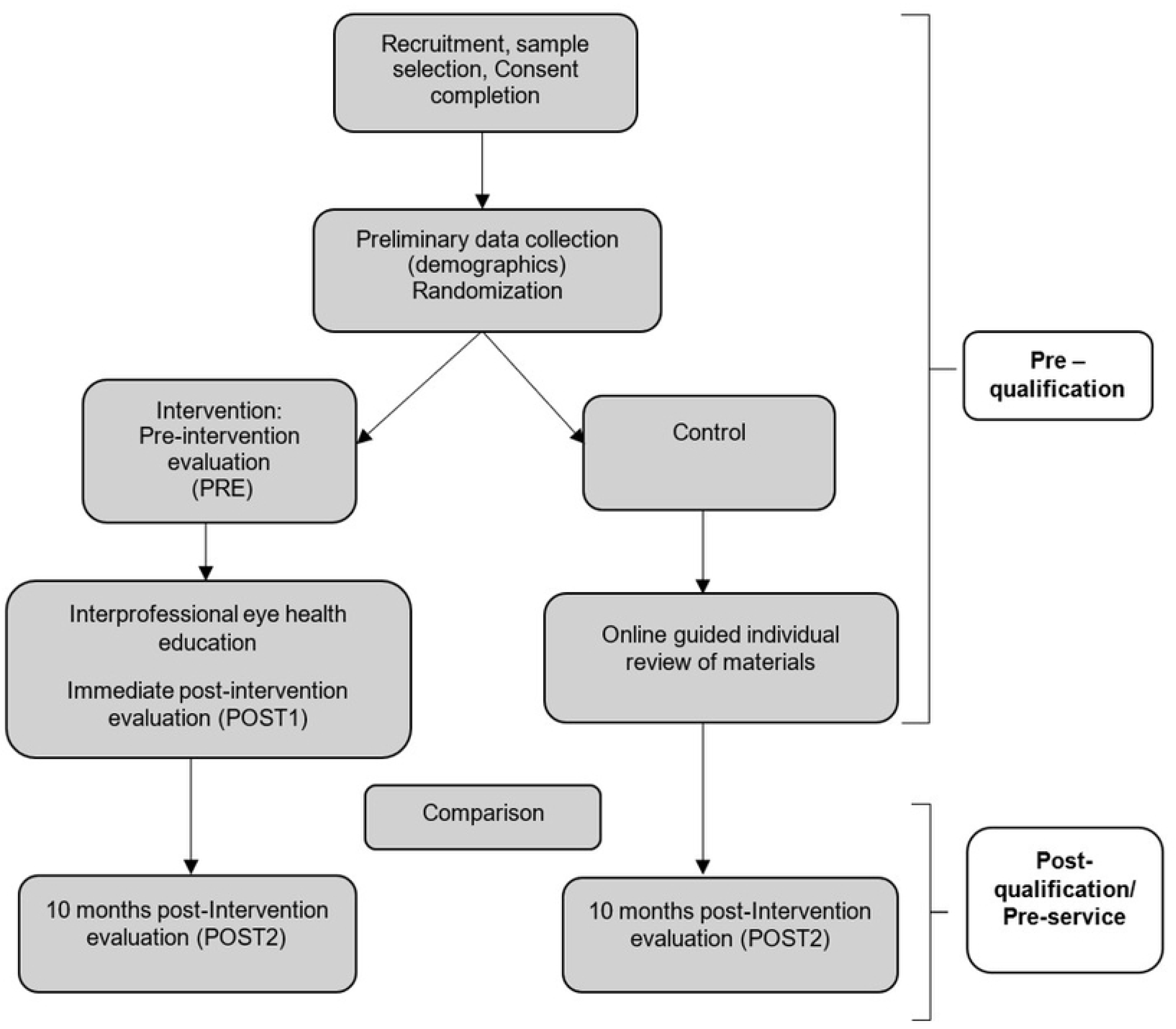
Illustration of the study flow and data collection points.

The knowledge assessment comprised 10 multiple-choice questions divided into two sections: Interprofessional collaborative practice (IPC) knowledge and fundamental eye health knowledge. Skills were assessed using a four station Objective Structured Clinical Examination (OSCE) to assess: quick holistic history taking for a patient with an eye problem, measuring distance vision, fundal reflex testing and, red eye management and collaborative eye care for people with diabetes and hypertension. The assessment tools went through a robust validation process, and the OSCE assessment was conducted by 7 trained interprofessional facilitators.^(30)^

Free text feedback was provided by the intervention group at the immediate post-training assessment (POST 1) via a short feedback form. The questions explored participants’ reflections on the training’s effectiveness and practical relevance. Specifically, they sought to identify aspects of the training perceived as most useful for their learning, factors that could facilitate the application of acquired knowledge in practice, and barriers that might hinder such implementation. Additionally, the open-ended question invited further suggestions or comments to improve the training’s structure, content, and overall impact. The data collection timeline for both intervention and control groups is outlined in (Figure2).

### Analysis

The Canadian Interprofessional Health Collaborative competencies^(32)^ and the Kirkpatrick model of training evaluation adapted by Barr^(29)^ were used to theoretically frame the analysis. Paired t-tests were conducted to compare pre and post-means of the overall scores for knowledge and skills. Non-parametric data were analysed using the Wilcoxon signed rank test and Kruskal-Wallis test. Post-hoc tests were conducted to compare the intervention and control group knowledge and skills 10 months after the intervention was implemented. Ethical approval from the Rwanda National Ethics Committee (RNEC) was granted (318/RNEC/ 2021).

A mixture of theoretical and reflexive thematic analysis^(33)^ was used for the analysis of qualitative data. GFK led the qualitative analysis guided by Braun and Clarke’s^(34)^ guidelines on thematic analysis. Coding of data was initiated through data familiarization, followed by iterative and robust calibration of the codebooks, through group coding of 10 pieces of data by each of the co-authors (VO, AB, AL), coding 10 transcripts independently. An inter-rater reliability analysis was calculated to allow discussion for consensus and final codebook generation. Each reviewer was allowed to add codes that captured information not covered by the initial codebook drafted by GFK. GFK used the final codebooks to code all the remaining qualitative data. Following the completion of coding, GFK developed themes that were refined through discussion with VO, AB, AL.

## Results

Among the participants randomised to the intervention group (n=180), a total of 161 (89.4%) attended the training, of which 113 (70.2%) attended the follow-up at 10 months post-intervention (POST2). Of the 100 participants randomised to the control group, 90 (90%) attended assessments at 10 months post-intervention (POST2).

Male and female participants were equally represented in both the intervention and control groups. The mean age for participants in the intervention and control groups was 25.5 (SD= 3.4) and 24 (SD=1.5)SD=3.4) and 24 (SD=1.5), respectively. The sample description including demographic details of the participants in each profession are detailed in table 1 and table 2.

**Table 1:**
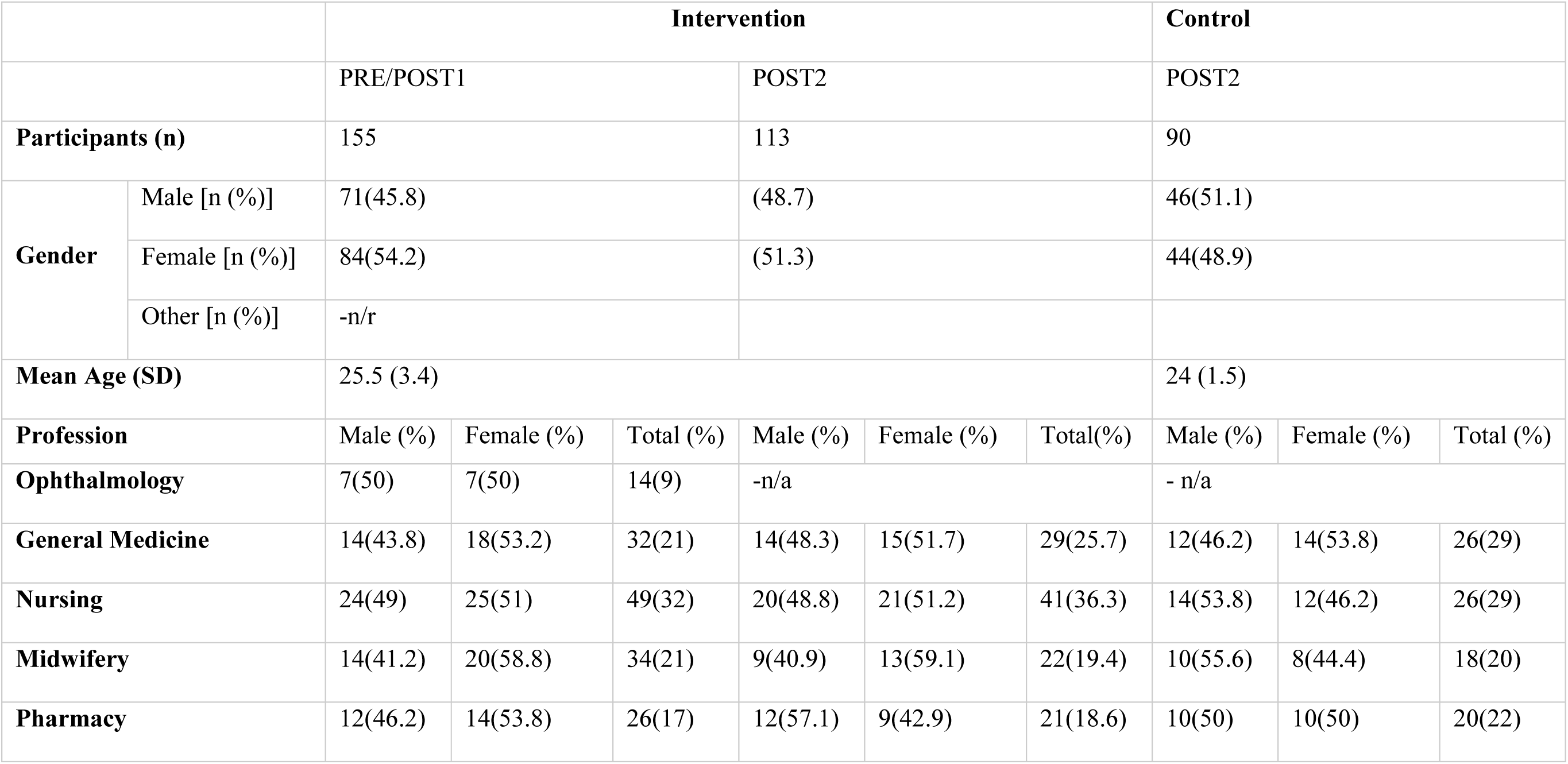
Demographic presentation of the sample of participants who attended the knowledge assessment.

**Table 2:**
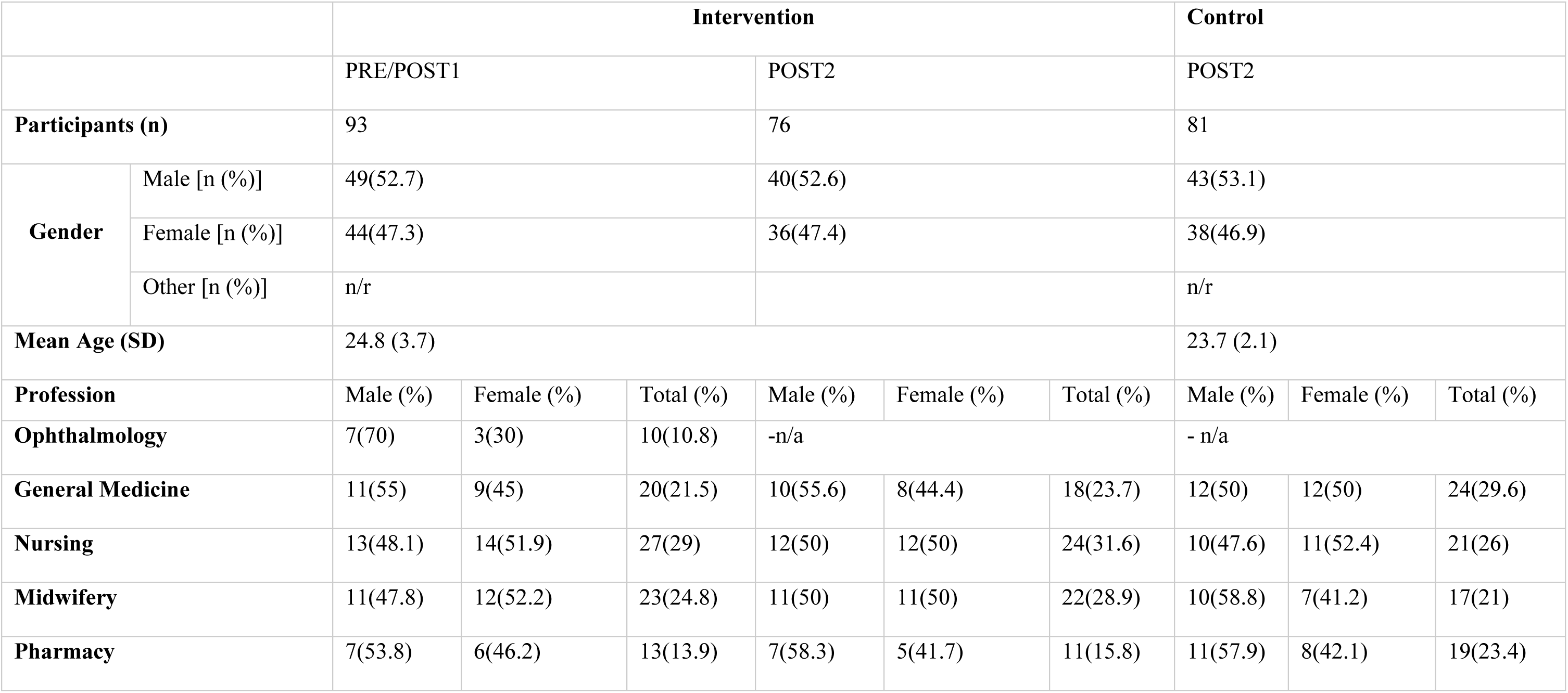
Demographic presentation of the sample of participants who attended the skills assessment.

|  |  | Intervention |  |  |  |  |  | Control |  |  |
| --- | --- | --- | --- | --- | --- | --- | --- | --- | --- | --- |
|  |  | PRE/POST1 |  |  | POST2 |  |  | POST2 |  |  |
| Participants (n) |  | 93 |  |  | 76 |  |  | 81 |  |  |
| Gender | Male [n (%)] | 49(52.7) |  |  | 40(52.6) |  |  | 43(53.1) |  |  |
|  | Female [n (%)] | 44(47.3) |  |  | 36(47.4) |  |  | 38(46.9) |  |  |
|  | Other [n (%)] | n/r |  |  |  |  |  | n/r |  |  |
| Mean Age (SD) |  | 24.8 (3.7) |  |  |  |  |  | 23.7 (2.1) |  |  |
| Profession |  | Male (%) | Female (%) | Total (%) | Male (%) | Female (%) | Total (%) | Male (%) | Female (%) | Total (%) |
| Ophthalmology |  | 7(70) | 3(30) | 10(10.8) | -n/a |  |  | - n/a |  |  |
| General Medicine |  | 11(55) | 9(45) | 20(21.5) | 10(55.6) | 8(44.4) | 18(23.7) | 12(50) | 12(50) | 24(29.6) |
| Nursing |  | 13(48.1) | 14(51.9) | 27(29) | 12(50) | 12(50) | 24(31.6) | 10(47.6) | 11(52.4) | 21(26) |
| Midwifery |  | 11(47.8) | 12(52.2) | 23(24.8) | 11(50) | 11(50) | 22(28.9) | 10(58.8) | 7(41.2) | 17(21) |
| Pharmacy |  | 7(53.8) | 6(46.2) | 13(13.9) | 7(58.3) | 5(41.7) | 11(15.8) | 11(57.9) | 8(42.1) | 19(23.4) |

### Knowledge assessment results PRE/ POST1/ POST2

155/161 intervention participants complete the pre- and post-intervention knowledge assessment, which gave a response rate of 96.2% of those who attended the training.

Mean IPC knowledge scores increased by an average of 60.6% pre- to post-training (SD = 30.8, Z = 10.55, p < 0.001, Wilcoxon signed-rank test). The increase in mean scores for fundamental eye health knowledge was 57.8% (SD=27.9, Z=10.61 p<0.001). Overall knowledge scores (combining IPC and fundamental eye health knowledge) increased by 58.9% (SD=20.8, Z=10.82 p<0.001).

There was no significant variation in PRE / POST1 differences in IPC knowledge between professional groups or gender groups. Ophthalmic clinical officers (OCOs) demonstrated advanced knowledge in fundamental eye health (PRE=82.1%, POST=96.4 %), which was also reflected in their overall knowledge score at the POST1 level, where they scored significantly higher than other professions. Data showed that for every 100 questions on fundamental eye health, OCO students scored an average of 73.2 points [H (4) = 73.2, p < 0.001] more than nursing students, for example. OCO students ended up with 57.7 [*H* (4) =57.7, p<0.001] more points in the overall knowledge assessment after the intervention compared to nursing students. OCOs also gained knowledge on fundamental eye health (mean difference= 14.3%, p<0.01) during the training intervention. There were no significant differences in eye health knowledge acquisition by gender.

The follow-up conducted at 10 months post-intervention (POST2) was completed by 113 intervention participants, 77.9% of the participants who attended the pre- and immediate post-intervention (POST1) sessions, excluding OCOs who were not invited to POST2. There was no statistically significant gender-related difference in attendance.

At 10 months after the intervention, the overall knowledge acquired during the intervention remained above the pre-intervention levels based on Wilcoxon signed rank test results (Figure 3) with intervention participants scoring an average of 75% (SD= 14.8) p< 0.001. However, compared to the scoring levels at POST1, there was a significant decrease of 13.3%(SD=19.6) p<0.01 in the average eye health knowledge score, while no significant decrease was recorded on IPC knowledge over the 10 months. A reduction in average score of 8.1% (SD=15.6) p<0.05 on the overall knowledge was recorded at 10 months (POST2).

**Figure 3:**
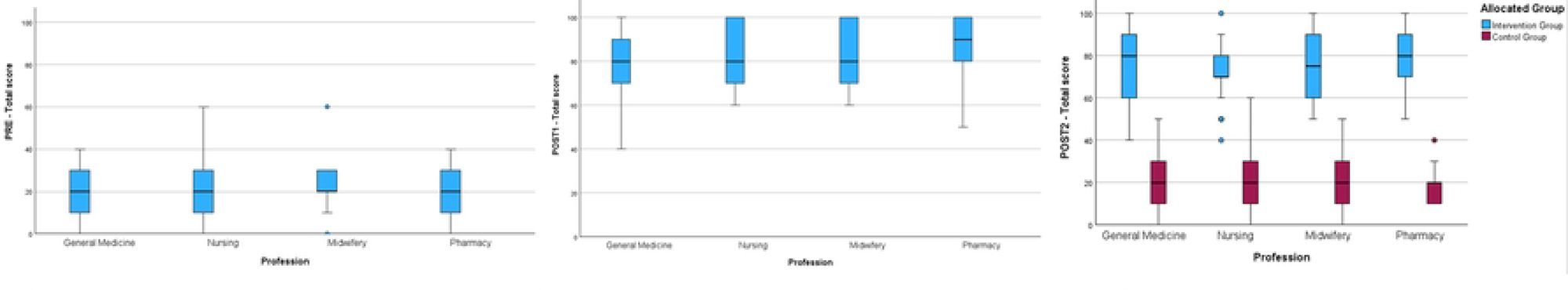
Boxplot graph comparing means of the overall knowlec.

There was no statistically significant difference in knowledge scores between professions or genders at 10 months follow-up based on a Dunn’s test conducted as part of a Kruskal-Wallis post hoc test.

There was no significant difference between the intervention group’s PRE overall knowledge scores and the control group’s POST2 overall knowledge scores (Z=−0.397, P=0.691). ANOVA analysis showed significant differences in IPC and fundamental eye health knowledge scores between the intervention and control group, respectively, at the POST2 timepoint [F(1, 201) = 521.10, p < 0.001 and F(1, 201) = 344.79, p < 0.001]. (Figure 3) Similarly, a t-test indicated a higher overall knowledge in the intervention group compared to control across all professions (score difference: 54.1%, SE= 2.0, df (201) = 27.3, P<0.001).

### Skills assessment results

Pre and POST1 intervention skills assessments were completed by 93 intervention group participants, resulting in a response rate of 57.7%. Of the participants who attended the pre and POST1 intervention skills assessment, 52.7% were male and the rest were female.

There was a significant overall skill acquisition after the intervention, with a mean difference in skills score pre and post-intervention of 49.7% (SD=14, Z= −8.382, P<0.001, Wilcoxon signed rank test). The greatest improvement in skills score was observed on the red reflex and visual acuity assessments, where the mean score difference was 59.5% (SD=21.4, Z=−8.363, P<0.001) and 55.4% (SD=19, Z=−8.409, P<0.001) respectively. No difference by gender or profession group was recorded.

A total of 76 intervention participants attended the 10-month follow-up skills assessment (POST2), and this made up 91.5% of all participants who attended the POST1 assessment excluding the OCOs. Among the participants who attended POST2 skills assessment, 52.6% were male and the remaining were female.

There was a significant decline in skills scores POST1 - POST2 in the intervention group for holistic ocular history taking [9.8%, SD=24.2, t(75)= 3.5, p<0.01], visual acuity assessment [9.6%, SD= 16.5, t(75)= 5.1, P<0.001], red reflex assessment [12%, SD= 17.6, t(75)= 5.937, p<0.001], and in overall skills scores [8.9%, SD= 10.5, t(75)= 7.4, p<0.001], but not for collaborative management of eye condition skills (Figure 4). Decline in skills was generally similar across professions in the intervention group, except for the overall skills score, where midwifery participants skills scores declined less than general medicine participants (9%, CI= 0.4 – 17.5, P<0.03). Further analysis showed that midwifery and pharmacy participants had less decline in overall skills scores at POST2 with 4.4% (CI= 0.7 – 7.9, P<0.017) and 4.8% (CI= −2.3 – 11.9, P<0.163) respectively compared to general medicine and nursing participants who had 13.4% (CI=8.5 – 18.3, p<0.001) and 11.8% (CI= 7.2 – 16.4, P<0.001) respectively. Gender was included in the multivariate analysis and the findings showed no effect of gender on the skills decline results. The recorded decline in overall skills scores did not reach the pre-intervention level (t=−36.2, df=75, P<0.001) in the intervention group.

**Figure 4:**
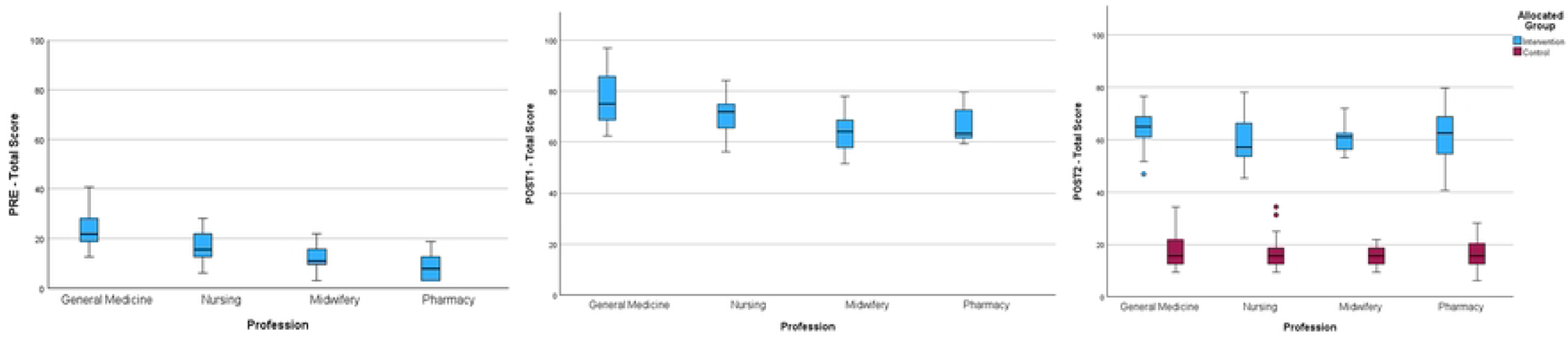
Boxplot graph comparing mean overall skill scores at P.

There was no difference in overall skills between the intervention group at the PRE timepoint and the control group at POST2 (t= −1.087, df=155, p=0.279). The intervention group scored significantly higher in overall skills than the control group at POST2 however, with mean difference of 44% (SE= 1.1, t (155) = 38.7, p<0.001). (Figure 4) The difference in skills score between the intervention and control group at POST2 was significant for: holistic ocular history Taking [ F (1, 155) = 59.044, p < 0.001], visual acuity assessment [F (1, 155) = 734.640, p < 0.001], fundal reflex assessment [F(1, 155) = 732.548, p < 0.001], and collaborative management of eye conditions [F(1, 155) = 282.742, p < 0.001].Qualitative findings

At POST1, 87(56.1%) of intervention participants provided free text comments about their experiences of the IPEHE. Demographic information of participants providing qualitative feedback can be found in table 3.

**Table 3:** Demographic description of participants who provided free-text feedback.

**Intervention Group (PRE/POST1)**
| Participants (n) |  | 87 |  |
| --- | --- | --- | --- |
| Gender | Male [n (%)] | 33(46) |  |
|  | Female [n (%)] | 47(54) |  |
|  | Other [n (%)] | n/r |  |
| Mean Age (SD) |  | 25.5 (3.4) |  |
| Profession | Male (%) | Female (%) | Total (%) |
| Ophthalmology | 2(40) | 3(60) | 5(5.7) |
| General Medicine | 8(36.3) | 14(63.7) | 22(25.3) |
| Nursing | 13(52) | 12(48) | 25(28.7) |
| Midwifery | 7(43.8) | 9(56.2) | 16(18.4) |
| Pharmacy | 9(47.4) | 10(52.6) | 19(21.9) |

Key themes that emerged from the thematic analysis are presented based on the University of British Columbia model^(35)^ of IPE (exposure, immersion, and mastery). Themes included: IPE link to better patient outcomes, eye health care, active interprofessional group work, resource acquisition and appreciation, increase in common knowledge and skills for IPC, hierarchy amongst professions, lack of awareness in colleagues, and resource issues (Figure 5).

**Figure 5:**
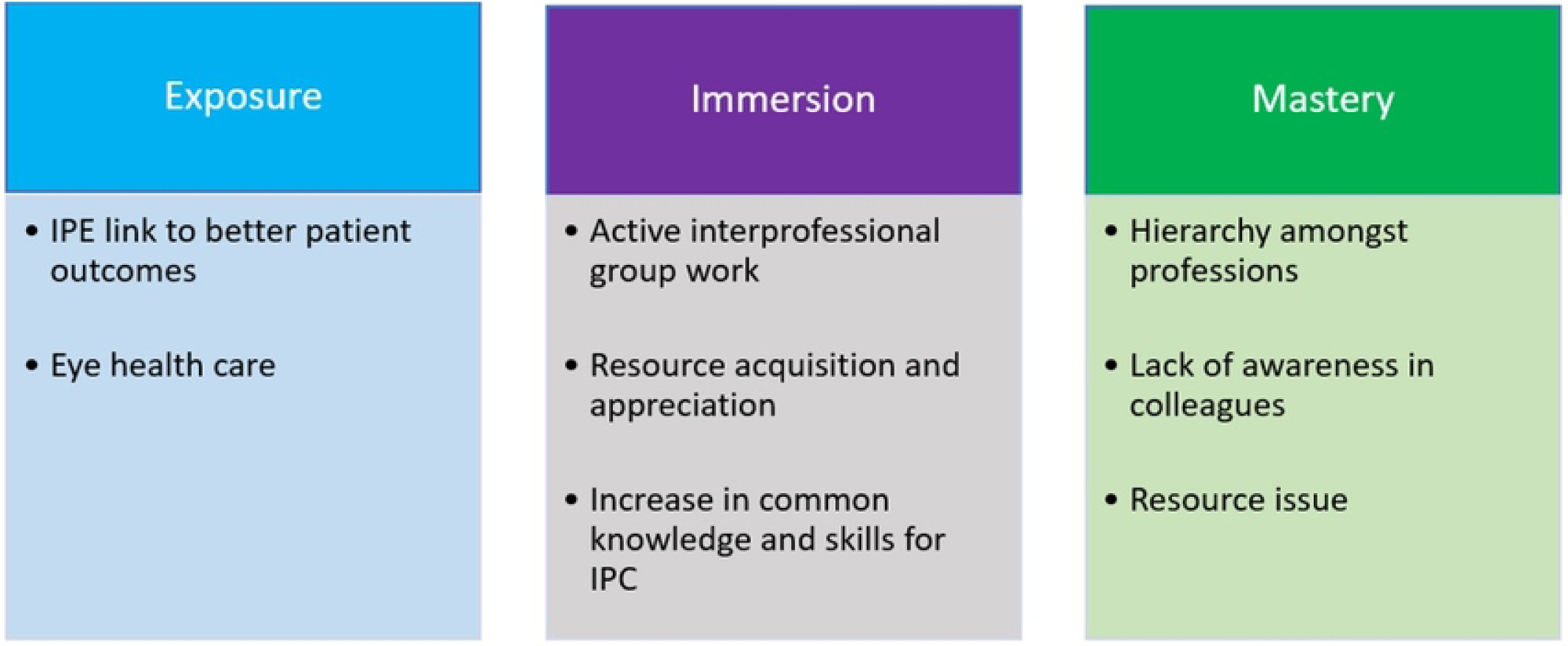
The University of British Columbia model (35) of IPE anc.

In relation to exposure and the theme of improving patient outcomes, participants shared insights on how the training equipped them with an understanding of their roles in healthcare, the role of colleagues from other professions, and how IPE brings healthcare providers to collaborate towards the same goal of getting patients back to functioning and participation in the community. The ICF framework^(36)^ was among the crucial components of the training. A holistic approach to patient care, functioning, and participation are main components in the ICF framework, and participants reflected those components among the key take-home messages from the training. The latter is illustrated in selected quotes below:

> “My useful part of the training was interprofessional collaboration how you collaborate to achieve better outcome to the clients (helping them return to their jobs)” – (Transcript 11, Nursing)

> “The useful part was first, a good understanding of everyone’s role in holistic approach to patient care.” – (Transcript 48, Pharmacy)

There was clear appreciation of the IPE approach, interactive learning and active participation that occurred as part of learning, and this was captured under the immersion theme of the UBC model. Participants reported the IPE approach using the Arclight package positively contributed to their learning. One participant shared:

> “The useful part of training was that we as trainees were engaged in this training and learnt together […with fellows…] from ophthalmology.” – (Transcript 42, General Medicine)

> “My most useful part was practical sessions, sharing ideas in group and using videos to learn.” – (Transcript 82, Ophthalmology)

> “ […] learning from ophtalmology how to use the Arclight and check for eye problems was very important to me […]” (Transcript 69, pharmacy)

On the mastery theme, participants shared concerns about the transfer of learning into practice, where issues such as hierarchy of professions and integrating collaborative eye health in existing health facility structures were among the points shared.

> “As a student, it can be hard to convince the doctors for example about interprofessional collaboration.” – (Transcript 55, General Medicine)

> “[…] behaviours of some health providers in hospitals who might resist to change from routines.” – (Transcript 10, Nursing)

## Discussion

Beyond the initial pilot studies exploring the feasibility of using the Arclight package for interprofessional education^(6),(15)^, this is the first comprehensive study to demonstrate that IPE can effectively enhance collaborative eye health skills in the short term among healthcare professionals in Sub-Saharan Africa. This work demonstrates that participants acquired fundamental ophthalmology and collaborative eye health knowledge through exposure to the IPEHE intervention and that IPE, as a means to support delivery, contributed to this change. The acquisition of knowledge is consistent with findings from a previous BEME systematic review^(37)^ that shows IPE can improve healthcare knowledge in both pre-qualification training and in practice. A cluster randomised controlled trial conducted in Rwanda^(38)^ also confirmed that IPE with the additional use of the ICF framework can improve knowledge of interprofessional practice at least in the short term^(39)^. The findings from this study and previous evidence on the effects of an IPE pilot study in delivering eye health training by O’Carroll and colleagues^(15)^, confirm that IPE can have a significant effect on shorter term eye health knowledge acquisition. This study also adds to the existing body of literature^(10),(15),(37),(40)^ demonstrating that IPE can contribute to healthcare knowledge and skills acquisition.

The qualitative data support the quantitative findings regarding knowledge and skill acquisition and describe how the IPE approach played a key role in that process. The learning steps described by participants were similar to the evidence reported in a systematic review by Aldriwesh et al^(41)^. The systematic review highlighted positive learning through interprofessional exposure, immersion, and mastery in undergraduate health professions education. The specialty areas covered by that review included: acute care, interprofessional communication, patient-centered practice, and creative and research skills. According to the systematic review by Aldriwesh and colleagues, there is a tendency for IPE research in undergraduate education to only focus on one step of the UBC model^(35)^ with IP exposure being the most common whilst immersion and mastery are least explored^(41)^. This study, however, explored how all three steps contributed not only to learning but also to impact on future mastery within healthcare facilities. Findings suggest that IPE did contribute to collaborative eye health knowledge and skills acquisition in undergraduate health professions education in Rwanda. Participants had time to focus on collaborative practice and eye health theory (knowledge), which was the main focus of the exposure step.^(35),(41)^ They went on to practice and learn with and from each other about eye health while putting into action the principles of collaborative practice (immersion), before considering the future mastery step in practice.

Even though the ICF framework^(11)^ was mainly applied to ensure participants had a common goal and language in the learning process, the framework also helped participants to better understand IPC competencies. This was reflected in the qualitative analysis. These findings complement the evidence^(11),(38),(42)^ that the ICF framework can be a catalyst for Interprofessional Education and Collaborative Practice (IPECP).

Crucially, the findings also showed collaborative eye health knowledge and skills retention in the longer term (10 months following IPEHE training) was present. There is limited evidence on the longer-term outcomes of IPE in SSA and globally; therefore, this study adds significantly to the current evidence.^(10)^ This study demonstrated limited decline in knowledge scores POST1/POST2 which did not go below 70% for both the overall knowledge and skills. This is usually considered high performance under the Rwandan education system.^(43)^ Skills scores declined slightly more POST1/POST2 but did not drop below 60%. Furthermore, both knowledge and skills scores were significantly higher in the intervention group compared with the control group at the POST2 timepoint.

Except for the collaborative management of eye conditions skill, there was a small but significant decline in skills acquired after 10 months, but this was still well above baseline. Decline in skills could be due to the time gap graduates spent without clinical practice in hospitals between graduating from university and entering practice. Additionally, as reflected in the qualitative data relating to mastery, intervention participants had anticipated challenges that were likely to hinder the transfer of learning in practice, and these may have contributed to a decline of acquired knowledge and skills. It is likely this gap of 2 to 3 months before starting practice with associated lack of opportunities to practice skills increases the likelihood of loss of skills consistent with previous studies by Gozie et al^(44)^ with refresher training effectively contributing to knowledge and skills retention as well as an increase in confidence in learners ^(40, 47)^. .

### Strengths and limitations

There are multiple strengths that contributed to the robust nature and transferability of the findings discussed in this study.

Key theoretical frameworks in the evaluation of IPE outcomes and impact underpinned this mixed-methods study, enhancing transferability to other contexts, including (but not exclusive to) other SSA countries. The study team was equipped with diverse expertise, comprising an ophthalmologist and global eye health specialist, health professions education and IPE specialists, and mixed-methods research specialists relevant to this study. This expertise not only contributed to vigorous study planning and implementation but also to the robust thematic analysis, specifically in refining the codebook and themes. Materials and resources used in the intervention and data collection were validated and piloted before data collection, strengthening the study. Also, where possible the confounding factors were equally distributed between the intervention and control groups using randomisation to minimise measurement errors.

Use of the Arclight package, a low-cost, portable diagnostic tool, alongside the ICF framework in eye health education, represents an innovative approach that addresses the scarcity of resources in low-income settings and demonstrates a practical application of frugal innovation in healthcare education which is replicable and scalable in other LMICs.

The study recruited non-ophthalmology participants with no previous ophthalmology training other than what was covered in their respective curricula. The low baseline in ophthalmology knowledge and skills contributed to the strength of the study in being able to measure changes from the eye care focussed intervention outcomes.

The eye health training program was well received, with participants showing consistent enthusiasm and high response rates throughout the study. Student retention was excellent, reflecting strong engagement and the overall acceptability of the intervention. This high level of continued participation enabled meaningful comparisons across different follow-up phases.

One of the factors that was not controlled for, but could have influenced the results, was the students’ cognitive performance in their respective departments at the University of Rwanda. Students who were the best performers in their respective classes may have engaged more and performed better in the IPEHE intervention and assessments. Stratification based on previous performance could help mitigate this source of variability and bias.

### Conclusion

This paper explored the short- and longer-term outcomes of an IPEHE intervention on collaborative eye health knowledge and skills amongst medical and health sciences students at the University of Rwanda. A strong and statistically significant improvement in collaborative eye health knowledge and skills attributed to the IPEHE intervention was found. This study is the first to show that this improvement in knowledge and skills is retained for up to 10 months following the IPEHE intervention. It was also noted that participants perceived that the IPE approach used to deliver the intervention positively contributed to learning and knowledge and skills acquisition.

There are implications from this study that are highly relevant to higher education institutions and educators in Rwanda and other LMICs faced with maximising the potential of their health workforce while at the same time aspiring to manage a large burden of eye disease. The findings from this study contribute to the existing body of knowledge by describing evidence for the feasibility of equipping HCPs not only with fundamental eye health knowledge, skills, and diagnostic tools. The Arclight ophthalmoscope does not necessarily need expert ophthalmic skill and contributed to equipping HCPs with key collaborative practice competencies during the pre-qualification phase with longer term retention of knowledge and skills.

With the adoption of the pre-qualification educational approach described in this study, training institutions can provide a collaborative practice-ready workforce capable of offering quality care to patients.

## Data Availability

The data underlying the results presented in the study are available from the University of St-Andrews Repository (https://pure.st-andrews.ac.uk/) or upon written request to the corresponding author.

## Reference

1. Bourne R, Adelson J, Flaxman S, Briant PS, Taylor HR, Casson RJ, et al. Trends in Prevalence of Blindness and Distance and Near Vision Impairment Over 30 Years and Contribution to the Global Burden of Disease in 2020. SSRN Electron J [Internet]. 2020 Aug 6 [cited 2021 Feb 2]; Available from: https://papers.ssrn.com/abstract=3582742

2. WHO. World report on vision. Vol. 214, World health Organization. 2019.

3. Humuza J, Uhawenimana TC, Ndayambaje JB, Matutina SU, Kananura T, Mungarulire J, et al. Evaluating the access and utilization of eyecare services after the adoption of eyecare PBF and its financial sustainability: the case of Rwanda. BMC Health Serv Res [Internet]. 2025 Dec 1 [cited 2025 Jul 22];25(1):1–13. Available from: https://bmchealthservres.biomedcentral.com/articles/10.1186/s12913-025-12776-9

4. Bright T, Kuper H, Macleod D, Musendo D, Irunga P, Yip JLY. Population need for primary eye care in Rwanda: A national survey. PLoS One. 2018;13(5):1–15.

5. Bourne RRA, Flaxman SR, Braithwaite T, Cicinelli M V., Das A, Jonas JB, et al. Magnitude, temporal trends, and projections of the global prevalence of blindness and distance and near vision impairment: a systematic review and meta-analysis. Lancet Glob Heal. 2017;5(9):e888–97.

6. O’Carroll V, Sagahutu JB, Ndayambaje D, Kayiranga D, Fiston Kitema G, Rujeni N, et al. Evaluation of a pilot interprofessional Arclight^TM^ workshop for healthcare students in Rwanda: promoting collaborative practice in eye health. J Interprof Care [Internet]. 2020;00(00):1–4. Available from: 10.1080/13561820.2020.1782356

7. WHO. Framework for action on interprofessional education & collaborative practice [Internet]. 2010 [cited 2021 Dec 3]. Available from: https://www.who.int/publications/i/item/framework-for-action-on-interprofessional-education-collaborative-practice

8. CAIPE. Statement of purpose. 2016; Available from: https://www.caipe.org/resource/CAIPE-Statement-of-Purpose-2016.pdf

9. Botma Y, Snyman S. Africa Interprofessional Education Network (AfrIPEN). J Interprof Care [Internet]. 2019;33(3):274–6. Available from: 10.1080/13561820.2019.1605236

10. Kitema GF, Laidlaw A, O’Carroll V, Sagahutu JB, Blaikie A. The status and outcomes of interprofessional health education in sub-Saharan Africa: A systematic review. J Interprof Care [Internet]. 2023 [cited 2023 Feb 27]; Available from: https://pubmed.ncbi.nlm.nih.gov/36739570/

11. Snyman S, Von Pressentin KB, Clarke M. International Classification of Functioning, Disability and Health: Catalyst for interprofessional education and collaborative practice. J Interprof Care. 2015;29(4):313–9.

12. MOH. REPUBLIC OF RWANDA National Strategic Plan for Eye Health. 2018;(December).

13. Mathenge C. Implementing an integrated primary eye care programme in Rwanda. Community Eye Heal [Internet]. 2022 [cited 2025 Apr 20];34(113):79. Available from: https://pmc.ncbi.nlm.nih.gov/articles/PMC9412126/

14. Reeves S, Perrier L, Goldman J, Freeth D, Zwarenstein M. Interprofessional education : effects on professional practice and healthcare outcomes (update) (Review) SUMMARY OF FINDINGS FOR THE MAIN COMPARISON. Cochrane Database Syst Rev. 2018;3(3).

15. Carroll VO, Kousha O, Kitema GF, Baptiste J, Denys N, Kayiranga D, et al. Interprofessional Arclight eye health workshop : impact on students ’ clinical identification and ophthalmic skills. J Interprof Care [Internet]. 2023;00(00):1–5. Available from: 10.1080/13561820.2023.2168257

16. Creswell JW. A concise introduction to mixed methods research. 2015.

17. Rwanda MOH. Ministry of Health Fourth Health Sector Strategic Plan. Minist Heal [Internet]. 2018;(July):1–104. Available from: http://moh.gov.rw/fileadmin/templates/Docs/FINALH_2-1.pdf

18. CONSORT. Consort - Welcome to the CONSORT Website [Internet]. [cited 2023 Jan 5]. Available from: https://www.consort-statement.org/

19. Moher D, Hopewell S, Schulz KF, Montori V, Gøtzsche PC, Devereaux PJ, et al. CONSORT 2010 explanation and elaboration: updated guidelines for reporting parallel group randomised trials. BMJ. 2010;340.

20. Universität Düsseldorf: G*Power [Internet]. [cited 2022 Nov 29]. Available from: https://www.psychologie.hhu.de/arbeitsgruppen/allgemeine-psychologie-und-arbeitspsychologie/gpower

21. Bruce CL, Juszczak E, Ogollah R, Partlett C, Montgomery A. A systematic review of randomisation method use in RCTs and association of trial design characteristics with method selection. BMC Med Res Methodol [Internet]. 2022;22(1):1–9. Available from: 10.1186/s12874-022-01786-4

22. Randomizer. Research Randomizer [Internet]. [cited 2022 Nov 26]. Available from: https://www.randomizer.org/

23. Barr H, Low H. Principles of Interprofessional Education. Cent Adv Interprofessional Educ. 2011;1–4.

24. Arclight. Education and training | Arclight Project [Internet]. [cited 2023 Feb 25]. Available from: https://medicine.st-andrews.ac.uk/arclight/devices/arclight/

25. Dooley E, Kousha O, Msosa J, Ndaule E, Abraham C, Parr J, et al. Comparative evaluation of a low cost direct ophthalmoscope (Arclight) for red reflex assessment among healthcare workers in Malawi. BMJ Innov. 2020;(figure 1):1–4.

26. Blaikie A, Sandford-Smith J, Tuteja SY, Williams CD, O’Callaghan C. Arclight: A pocket ophthalmoscope for the 21st century. BMJ. 2016;355.

27. Blaikie AJ, Dutton GN. How to assess eyes and vision in infants and preschool children. BMJ. 2015;350.

28. WHO. Primary Eye Care training manual | WHO | Regional Office for Africa [Internet]. 2018 [cited 2021 Jan 17]. Available from: https://www.afro.who.int/publications/primary-eye-care-training-manual

29. Craig P, Hall S, Phillips C. Using the Freeth/Kirkpatrick model to evaluate interprofessional learning outcomes in a rural setting. Focus Heal Prof Educ A Multi-Professional J. 2016;17(1):84.

30. Kitema GF. Impact of interprofessional eye health education using the Arclight training package and the International Classification of Function Disability and Health (ICF) framework among health sciences students in Rwanda: a mixed methods study [Internet]. University of St-Andrews; 2024. Available from: 10.17630/sta/953

31. Forman D, Thistlethwaite J. Best practice in leading research and evaluation for interprofessional education and collaborative practice. Leading Research and Evaluation in Interprofessional Education and Collaborative Practice. 2016. 3–21 p.

32. Canadian Interprofessional Health Collaborative. Cihc [Internet]. A National Interprofessional Competency Framework. 2010. 1–32 p. Available from: http://www.cihc.ca/files/CIHC_IPCompetencies_Feb1210.pdf

33. Braun V, Clarke V. Using thematic analysis in psychology. Qual Res Psychol. 2006;3(2):77–101.

34. Braun V, Clarke V. Successful Qualitative Research: A Practical Guide for Beginners [Internet]. 1st ed. SAGE Publications. London: SAGE Publications; 2013 [cited 2023 Mar 28]. 400 p. Available from: https://www.researchgate.net/publication/256089360_Successful_Qualitative_Research_A_Practical_Guide_for_Beginners

35. Charles G, Bainbridge L, Gilbert J. The University of British Columbia model of interprofessional education. J Interprof Care. 2010;24(1):9–18.

36. Moran M, Bickford J, Barradell S, Scholten I. Embedding the International Classification of Functioning, Disability and Health in Health Professions Curricula to Enable Interprofessional Education and Collaborative Practice. J Med Educ Curric Dev. 2020;7:238212052093385.

37. Reeves S, Fletcher S, Barr H, Birch I, Boet S, Davies N, et al. A BEME systematic review of the effects of interprofessional education: BEME Guide No. 39. Med Teach. 2016;38(7):656–68.

38. Sagahutu JB, Kagwiza J, Cilliers F, Jelsma J. The impact of a training programme incorporating the conceptual framework of the International Classification of Functioning (ICF) on knowledge and attitudes regarding interprofessional practice in Rwandan health professionals: a cluster randomized contro. BMC Med Educ. 2021;21(1):1–13.

39. WHO. How to use the ICF - A Practical Manual for using the International Classification of Functioning, Disability and Health [Internet]. World health Organization. 2013 [cited 2023 Mar 1]. Available from: https://www.who.int/publications/m/item/how-to-use-the-icf---a-practical-manual-for-using-the-international-classification-of-functioning-disability-and-health

40. Namagembe A, Ssekabira U, Weaver MR, Blum N, Burnett S, Dorsey G, et al. Improved clinical and laboratory skills after team-based, malaria case management training of health care professionals in Uganda. 2012;1–10.

41. Aldriwesh MG, Alyousif SM, Alharbi NS. Undergraduate-level teaching and learning approaches for interprofessional education in the health professions: a systematic review. BMC Med Educ [Internet]. 2022;22(1):1–14. Available from: 10.1186/s12909-021-03073-0

42. Sagahutu JB, Kagwiza J, Cilliers F, Jelsma J. The impact of a training programme incorporating the conceptual framework of the International Classification of Functioning (ICF) on behaviour regarding interprofessional practice in Rwandan health professionals: A cluster randomized control trial. PLoS One [Internet]. 2020;15(2):1–17. Available from: 10.1371/journal.pone.0226247

43. scholaro. Grading System in Rwanda [Internet]. Scholaro Database. 2023 [cited 2023 Sep 18]. Available from: https://www.scholaro.com/db/countries/Rwanda/Grading-System

44. Offiah G, Ekpotu LP, Murphy S, Kane D, Gordon A, O’Sullivan M, et al. Evaluation of medical student retention of clinical skills following simulation training. BMC Med Educ. 2019;19(1):1–7.

